# Optimization of a thermal shaker-aided quaking-induced conversion platform for the rapid detection of prion seeding activity in human samples

**DOI:** 10.64898/2026.09.10.26362572

**Authors:** Yukiko Miyazaki, Takujiro Homma, Takehiro Nakagaki, Hanae Takatsuki, Tsuyoshi Mori, Ryuichiro Atarashi, Katsuya Satoh, Daisuke Ishibashi

**Affiliations:** Department of Microbiology and Immunology, Graduate School of Biomedical Sciences, Nagasaki University, Nagasaki, Japan; Department of Pharmacology, Graduate School of Medicine, Osaka Metropolitan University, Osaka, Japan; Division of Microbiology, Department of Infectious Diseases, Faculty of Medicine, University of Miyazaki, Japan; Department of Locomotive Rehabilitation Science, Graduate School of Biomedical Sciences, Nagasaki University, Nagasaki, Japan; Department of Immunological and Molecular Pharmacology, Faculty of Pharmaceutical Science, Fukuoka University, Fukuoka, Japan; Present address: Department of Protozoology, Institute of Tropical Medicine (NEKKEN), Nagasaki University, Nagasaki, Japan

## Abstract

Creutzfeldt–Jakob disease (CJD) is a rapidly progressive, fatal neurodegenerative disorder caused by the misfolding of the normal prion protein (PrP^C^) into its pathogenic form (PrP^Sc^). Therefore, effective clinical management and infection control require diagnostic assays that combine sub-attomole sensitivity with a turnaround time compatible with routine practice. While real-time quaking-induced conversion (RT-QuIC) offers excellent analytical performance, its protracted reaction time and dependence on plate readers that integrate high-speed shaking with continuous fluorescence detection restrict its widespread implementation.

Endpoint QuIC (EP-QuIC) is an established alternative approach that utilizes vigorous shaking in a ThermoMixer^®^ C and relies on a single endpoint fluorescence readout. In this study, we introduced minor modifications to assay conditions to enhance the speed, robustness, and reproducibility of EP-QuIC. The refined assay detected prion seeding activity in sporadic CJD brain samples within 6 h—and, in some cases, as early as 2 h. Unlike RT-QuIC, problematic recombinant PrP batches that impaired assay reliability in RT-QuIC—exhibiting delayed kinetics and spontaneous ThT fluorescence in unseeded reactions—did not produce such effects in EP-QuIC. Instead, they yielded strong, specific fluorescence signals that clearly identified sCJD samples, demonstrating improved batch tolerance and diagnostic efficiency of this approach. When applied to CSF, the EP-QuIC assay showed 100% sensitivity and specificity, highlighting its potential as a rapid, reliable diagnostic tool for prion diseases.

## Introduction

Prion diseases, including Creutzfeldt–Jakob disease (CJD) and bovine spongiform encephalopathy, are fatal and transmissible neurodegenerative disorders affecting humans and animals. Their pathogenesis is characterized by the aggregation of misfolded prion protein (PrP^Sc^) arising from the conformational conversion of the cellular prion protein (PrP^C^) [1]. Human prion diseases are divided into three major categories: sporadic CJD (sCJD), genetic prion diseases, and environmentally acquired prion diseases. Among these, sCJD is the most prevalent and is further classified based on the polymorphism at codon 129 of the *prion protein* (*Prnp*) gene (methionine homozygosity (MM), methionine/valine heterozygosity (MV), and valine homozygosity (VV)) and the proteinase K (PK) digestion pattern of PrP^Sc^ (types 1 and 2) [2]. These molecular subtypes are associated with distinct clinical symptoms and pathological features [2]. Currently, there is no cure for these diseases, and they inevitably progress to severe neurological decline and death.

A definitive diagnosis of sCJD requires the detection of PrP^Sc^ and characteristic histopathologic findings from the brain tissue, such as widespread spongiform degeneration, neuronal loss, gliosis, and PrP amyloid deposition [3]. Due to the invasive nature of brain biopsies, clinical antemortem diagnosis typically relies on non-invasive methods, including magnetic resonance imaging (MRI), electroencephalography (EEG), and cerebrospinal fluid (CSF) analysis [4–6]. In CSF analysis, elevated levels of 14-3-3 and total tau proteins are often used as supportive biomarkers [7]. However, the sensitivity of these tests varies widely across disease stages and sCJD subtypes [8,9], and they lack specificity for CJD [10], highlighting the urgent need for more accurate and reliable diagnostic methods.

To overcome diagnostic limitations, several cell-free amplification methods have been developed to detect trace amounts of PrP^Sc^ in tissues and fluids from individuals with CJD. One such method, protein misfolding cyclic amplification assay, promotes PrP conversion through repeated cycles of sonication, using either brain homogenates or recombinant PrP (recPrP) as substrates seeded with PrP^Sc^ from infected brains [11,12]. The quaking-induced conversion (QuIC) assay offers a simpler alternative by utilizing intermittent shaking to facilitate the conversion of recPrP substrates seeded with PrP^Sc^ [13]. Further advances led to the development of real-time QuIC (RT-QuIC), which enables real-time monitoring of amyloid fibril formation by detecting thioflavin T (ThT) fluorescence using a plate reader [14].

RT-QuIC enables ultrasensitive detection of previously undetectable PrP^Sc^ levels in CSF within 2–4 days, offering significantly higher specificity than conventional CSF biomarkers, such as 14-3-3 and total tau proteins [14]. Beyond its diagnostic value, RT-QuIC serves as a sensitive alternative to traditional animal-based bioassays for assessing prion infectivity, with the 50% seeding dose (SD_50_) derived from its endpoint serving as a reliable surrogate marker for prion titers [15, 16]. Furthermore, the assay has demonstrated widespread prion seeding activity outside the central nervous system in patients with sCJD [17] and has proven useful in testing the efficacy of prion decontamination protocols for surgical instruments [18]. More recently, RT-QuIC has been shown to be effective for detecting latent prion infection in cadavers, offering a potential means to mitigate occupational exposure risks for healthcare and anatomical personnel [19].

Despite its strengths, RT-QuIC has two major limitations. First, although it offers shorter lag times compared to other detection methods, the assay often requires extended incubation—sometimes exceeding 40–50 hours at 42°C—especially when detecting low PrP^Sc^ levels [20,21]. Second, batch-to-batch variability in the quality of in-house–produced recPrP might lead to poor reproducibility. Some studies have reported minimal batch-to-batch variability under tightly controlled conditions [22]. Reliable RT-QuIC performance demands strict control of parameters such as salt concentration, pH, and storage temperature. Even the different recPrP batches produced using similar protocols exhibit marked differences in their ability to support RT-QuIC assay, indicating that not all recPrP substrates are equally effective [23,24]. In addition, some commercial recPrP sources have failed to yield consistent results [25], highlighting the importance of careful batch selection. Furthermore, prolonged shaking can cause mechanical wear of plate readers and increase reagent waste, raising operational costs and limiting clinical adoption of RT-QuIC and elevate operational costs. Caughey et al. first reported using the ThermoMixer® R, a cold-top shaker, for QuIC with 263K scrapie hamster brain seed [13]. The same group later employed a heated plate shaker analogous to the ThermoMixer^®^ and reported shortened reaction times with 263K scrapie; however, they also reported increased fluorescence signals in negative control wells [23]. To address these limitations, an optimized method, termed endpoint QuIC (EP-QuIC), was developed by other investigators [26–28]. This method employs vigorous orbital shaking using ThermoMixer^®^ C and replaces real-time fluorescence monitoring with a single endpoint fluorescence measurement.

In the current study, we employed a thermal shaker-based approach, with slight modifications to the assay conditions, and directly compared its performance to that of standard RT-QuIC, aiming to determine whether the observed acceleration was an inherent feature of thermal mixers. The optimized protocol enabled highly sensitive detection of trace levels of prions in the CSF of patients with sCJD, reducing the total assay duration to just 4–6 h. Notably, the performance of the optimized assay remained consistent across different recPrP batches, marking a significant improvement in reproducibility. Furthermore, our optimized method offers uniform heat transfer and mixing, reduces mechanical stress on equipment, and minimizes reagent consumption. These enhancements make the assay faster, more reliable, and better suited for routine clinical diagnosis of prion diseases compared to traditional RT-QuIC.

## Materials and Methods

### Ethics Statement

The Ethics Committee of Nagasaki University Hospital approved the study protocol (Approval ID: 100428423), and the Japan CJD Surveillance Unit authorized the use of brain tissue. The study is registered with the University Hospital Medical Information Network under registration numbers UMIN000003301 and UMIN000038398. All samples were anonymized and analyzed in a blinded manner. Written informed consent was obtained from each participant or their legal guardian.

### Patients

Brain tissue samples were obtained from two patients with sCJD, confirmed by postmortem histopathological analysis. Both individuals were homozygous for methionine at codon 129 of *PRNP* and were classified as the MM1 subtype of sCJD. These two patients (P1 and P2) had been previously reported and analyzed using the RT-QuIC assay [16,17]. CSF samples were collected from ten additional patients (P3–P12) with a confirmed diagnosis of sCJD, as well as from ten control patients (N1–N10) diagnosed with other neurological disorders through the CJD Surveillance Committee in Japan. All patients were analyzed in this study. Demographic and clinical information for P1 and P2 is provided in Table 1, and that for P3–P12 and N1–N10 is provided in Supplementary Table 1.

**Table 1.** Clinical data and the SD_50_ concentrations at each reaction time in the sporadic CJD brain samples using E-QUIC and RT-QUIC.

| ID | Sex | Age range at death (years) | Final diagnosis |  | E-QUIC (Log SD<sub>50</sub>/g tissue) |  | RT-QUIC (Log SD<sub>50</sub>/g tissue) |  |
| --- | --- | --- | --- | --- | --- | --- | --- | --- |
|  |  |  | Codon 129 | WB type | 4 hr | 6 hr | 24 hr | 48 hr |
| P1 | Female | 66–70 | MM | 1 | 10.3 ± 0.35 | 10.3 ± 0.24 <sup>a</sup> | 9.7 ± 0.12 | 9.9 ± 0.12 |
| P2 | Male | 71–75 | MM | 1 | 10.3 ± 0.20 | 10.5 ± 0 | 8.8 ± 1.12 | 9.6 ± 0.42 |

All SD<sub>50</sub> values are presented as mean ± standard deviation (S.D.) and were calculated from at least three independent experiments.
MM: methionine homozygosity at codon 129 of *PRNP*.
WB type indicates the Western blotting pattern observed in each sample.
<sup>a</sup>: $P < 0.05$ versus the 24-h RT-QUIC value in Patient 1 (Student's *t*-test).

### Preparation of Brain Homogenates

Brain homogenates were prepared as described previously [17]. Briefly, samples from the frontal cortex of patient brains were weighed and homogenized in phosphate-buffered saline (PBS) to a final concentration of 10% (w/v). To avoid cross-contamination among samples, all steps were performed using single-use disposable tubes and beads, and each sample was processed on different days. The homogenates were centrifuged at 6000 rpm for 2 min and the supernatants was stored at −80°C.

### Expression and Purification of Recombinant PrP

The expression and purification of human recPrPs (residues 23–231 and 90–231) and hamster recPrP (residues 90–231) were performed as described previously [12,14]. After purification, aliquots of the recombinant proteins were stored at −80°C. For human recPrP 23–231, three independently prepared batches produced on different days (batches #1–3) were used in subsequent experiments.

### RT-QuIC

RT-QuIC was performed as described previously [14]. Unless otherwise specified, the reactions were conducted using human recPrP 23–231. Brain homogenates were serially diluted ten-fold in phosphate-buffered saline (PBS). The reaction mixture contained 500 mM sodium chloride (NaCl), 50 mM piperazine-N,N′-bis(2-ethanesulfonic acid) (PIPES) (pH 7.0), 1 mM ethylene diamine tetraacetic acid (EDTA), 0.001% sodium dodecyl sulfate (SDS), 10 µM thioflavin T (ThT), and 0.1 mg/mL of recPrP. Reactions were set up in clear-bottom 96-well plates (Greiner Bio-One, Frickenhausen, Germany). For brain homogenate samples, 10 µL of diluted homogenate was combined with 90 µL of reaction mixture per well. For CSF samples, 5 µL of diluted CSF was added to 95 µL of reaction mixture per well. Artificial CSF (A-CSF), containing 125 mM NaCl, 2.5 mM potassium chloride (KCl), 2 mM calcium chloride (CaCl_2_), 1 mM magnesium chloride (MgCl_2_), 0.2 ng/mL Bovine serum albumin (BSA), and 0.05% glucose, was used where indicated. All samples were run in quadruplicates. The plates were sealed and incubated at 37°C in an Infinite^®^ 200 PRO plate reader (Tecan, Zurich, Switzerland) with alternating cycles of 30 s of orbital shaking at maximum speed and 30 s of rest for a total of 48 h. ThT fluorescence was measured every 10 min using excitation and emission wavelengths of 445 and 485 nm, respectively. The wells were considered positive when fluorescence exceeded twice the mean of “no-seed” controls. Seeding activity was quantified as the 50% seeding dose (SD_50_) using the Spearman–Kärber method.

### Thermal Shaker-Aided QuIC (EP-QuIC)

The reaction mixture composition used for EP-QuIC was the same as that used for RT-QuIC. For negative controls, an equal volume of PBS was added to “no-seed” wells. The plates were clamped onto a ThermoMixer^®^ C (Eppendorf, Hamburg, Germany), which was set to 42°C or 55°C, and were subjected to alternating cycles of 60 s of shaking at 1500 rpm and 60 s of rest for the indicated duration. During incubation, a ThermoTop^®^ (Eppendorf, Hamburg, Germany) was placed on the thermoblock to ensure uniform temperature control and protect the samples from light exposure. After 2, 4, 6, and 20 h, the plates were removed from the ThermoMixer^®^ C, and a final relative fluorescence unit reading was obtained using a FLUOstar OMEGA (BMG LABTECH, Ortenberg, Germany) plate reader. Ample absorbance was measured at excitation and emission wavelengths of 445 and 485 nm, respectively. Each dilution was analyzed in quadruplicate.

### Statistical Analysis

All statistical analyses were performed using GraphPad Prism. Seeded reactions were compared with the corresponding “no-seed” controls at each time point using two-way analysis of variance with Dunnett’s multiple comparisons test. Data were expressed as means ± standard deviations (SDs).

## Results

### Prion seeding activity is rapidly detected using a thermomixer-based EP-QuIC

We investigated a previously established amplification assay EP-QuIC, with modified experimental conditions, to assess its efficacy as a faster and more robust alternative to RT-QuIC. The reactions were conducted at 42°C with high-speed circular shaking on an Eppendorf Thermomixer^®^ C, with human recPrP 23–231 as the substrate. These conditions differ from those used in a typical EP-QuIC protocol, which uses hamster recPrP 23–231, shaking at 900 rpm with 90-s shaking and 30-s rest cycles, and a different reaction platform [27]. For comparison, conventional RT-QuIC was performed on a Tecan plate reader with real-time ThT fluorescence measurement over 48 h.

Brain tissues from P1 and P2 were analyzed to evaluate reaction kinetics. Under EP-QuIC conditions, a marked increase in ThT fluorescence was detected as early as 2 h at higher seed concentrations (10^−6^ g), whereas lower concentrations (10^−7^ to 10^−10^ g) became detectable within 4–6 h (Fig. 1A). In contrast, RT-QuIC required approximately 24 h to detect seeding activity at comparable concentrations (Fig. 1B). No spontaneous signal increase was observed in unseeded controls even after 20 h, indicating high specificity of the assay under optimized conditions. A dose–response comparison showed that EP-QuIC consistently produced a higher percentage of positive replicates at 4 and 6 h compared to RT-QuIC at 24 or 48 h, particularly at low homogenate concentrations (10^−9^–10^−10^ g, Fig. 1C). Notably, EP-QuIC detected prion seeding activity in brain homogenates from both patients within 4 h, demonstrating greater sensitivity than RT-QuIC over longer durations. Quantitative analysis revealed that the SD_50_ value for P1 was 9.7 ± 0.12 after a 6-h EP-QuIC reaction, significantly lower than the SD_50_ value (10.3 ± 0.24) observed with RT-QuIC after 24 h, whereas no significant difference was found for P2 (Table 1). These results highlight the enhanced speed and sensitivity of EP-QuIC when applied to standard sCJD brain homogenates, without the need for additional sample processing.

**Figure 1.**
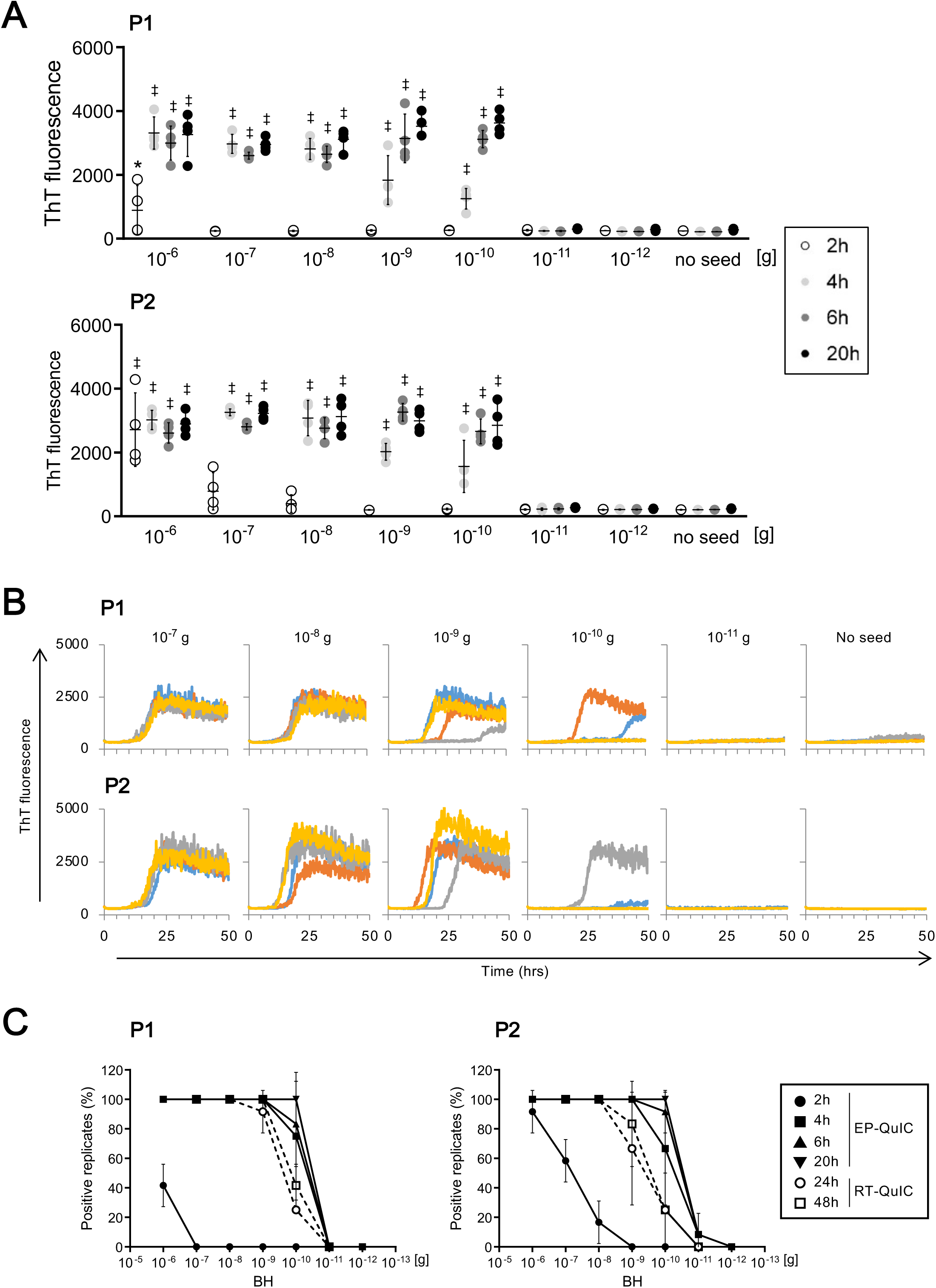
Comparison of endpoint-quaking-induced conversion (EP-QuIC) and real-time quaking-induced conversion (RT-QuIC) for prion detection in brain tissues. A) Thioflavin T (ThT) fluorescence in EP-QuIC reactions. Reactions were performed at 42°C using human recombinant prion protein (recPrP) 23–231 (batch #1) and brain homogenates from two patients with sporadic Creutzfeldt–Jakob disease (sCJD, P1 and P2) at concentrations of 10^−6^ to 10^−12^ g, along with “no-seed” controls. ThT fluorescence was measured at 2, 4, 6, and 20 h. \**P* < 0.05, ‡*p* < 0.001 *vs.* the “no-seed” controls at the corresponding time points. B) ThT fluorescence in RT-QuIC reactions. Reactions were performed at 42°C using human recPrP 23–231 (batch #1) and brain homogenates from two patients with sCJD (P1 and P2) at concentrations of 10^−7^ to 10^−11^ g, along with “no-seed” controls. ThT fluorescence was continuously monitored in real time for 48 h. C) Dose–response analysis showing the percentage of positive replicate reactions at each seed concentration for EP-QuIC (2, 4, 6, and 20 h) and RT-QuIC (24 and 48 h), calculated from the data shown in panels (A) and (B).

#### EP-QuIC performance is not affected by batch-to-batch variation in recPrP

Batch-to-batch variability in recPrP has been a major concern in conventional RT-QuIC, as certain batches tend to form spontaneous fibrils, resulting in poor reproducibility and potential false positives [23–25]. To investigate this issue, we examined two independently produced human recPrP batches (#2 and #3). Both batches exhibited spontaneous increases in ThT fluorescence in RT-QuIC reactions, even without prion seed addition (Fig. 2A). Although these batches were prepared using similar methods, they displayed notable differences in their ability to support prion seeding activity. RT-QuIC reactions with batch #2 produced positive results at brain homogenate concentrations of 10^−7^–10^−9^ g; however, the fluorescence increase was delayed, and maximal signal levels were often not achieved, indicating slower reaction kinetics. Batch #3 performed even more poorly, showing minimal or no amplification and failing to generate a detectable increase in ThT fluorescence, suggesting severely compromised reactivity. These results indicated that not all recPrP batches are equally suitable for RT-QuIC and that careful selection and validation of recPrP preparations are essential for reliable assay performance.

**Figure 2.**
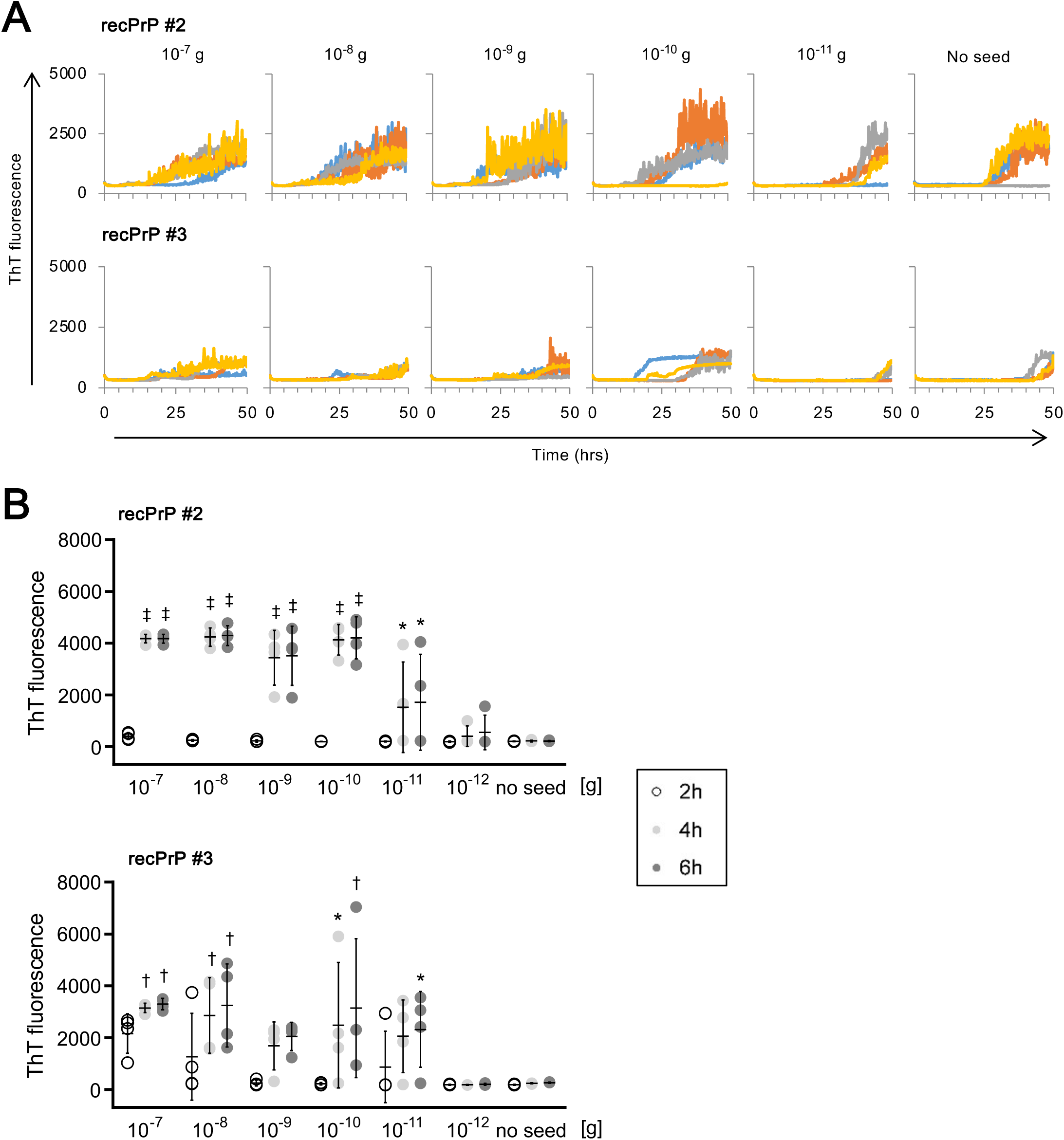
Robustness of EP-QuIC against recPrP batch variation. A) Effect of recPrP batch variability on RT-QuIC performance. Reactions were performed at 42°C using human recPrP 23–231 (batches #2 and #3) and brain homogenates from a patient with sCJD (P1) at concentrations of 10^−7^ to 10^−11^ g, along with “no-seed” controls. ThT fluorescence was continuously monitored in real time for 48 h. B) Effects of recPrP batch variability on EP-QuIC performance. Reactions were performed at 42°C using human recPrP 23–231 (batches #2 and #3) and brain homogenates from a patient with sCJD (P1) at concentrations of 10^−7^ to 10^−12^ g, along with “no-seed” controls. ThT fluorescence was measured at 2, 4, and 6 h. \**P* < 0.05, †*p* < 0.01, ‡*p* < 0.001 *vs.* the “no-seed” controls at the corresponding time point.

To determine whether EP-QuIC is similarly affected by batch-to-batch variation in recPrP, we tested the same two recPrP batches that had shown spontaneous aggregation in RT-QuIC. Using EP-QuIC, all replicate reactions with batch #2 were positive at brain homogenate concentrations of 10^−7^–10^−9^ g, whereas batch #3 also produced clear, distinguishable fluorescence signals at concentrations of 10^−7^–10^−8^ g. In both cases, increased ThT fluorescence was detectable within 6 hours (Fig. 2B). Furthermore, no signal increase was observed in unseeded control reactions for either batch (Fig. 2B). These findings demonstrated that EP-QuIC delivers consistent and reliable results across different recPrP batches and is not significantly affected by batch-to-batch variability.

#### EP-QuIC reaction conditions are optimized for improved sensitivity and specificity

Previous research has demonstrated that raising the reaction temperature to 55°C significantly boosts ThT fluorescence signals and reduces lag times compared to reactions at 42°C [23, 29–31]. To investigate whether higher temperatures would similarly enhance EP-QuIC performance, we conducted reactions at 55°C (Supplementary Fig. 1A). Under these conditions, maximum ThT fluorescence was achieved within 2–6 hours across all tested brain homogenate concentrations (10^−6^–10^−10^ g), with initial signal increases observed as early as 2 h. Although spontaneous aggregation was observed in batches #2 and #3 after 6 h at 55°C, the delayed onset of these signals made them clearly distinguishable from seeded reactions.

We next assessed the presence of SDS on EP-QuIC performance. SDS concentration has been reported to significantly affect the sensitivity and specificity of RT-QuIC, with 0.001% SDS generally enhancing reactivity [32] and even 0.002% SDS being effective as a positive regulator of reactions under certain conditions [29]. Higher SDS concentrations might inhibit the reaction or lead to false-positive results [32]. However, the effect of SDS on EP-QuIC has not yet been characterized. Thus, in the current study, we performed EP-QuIC using 0%, 0.001%, and 0.002% SDS (Supplementary Fig. 1B). Although detectable signals were observed in the absence of SDS, no fluorescence signals were detected at either 0.001% or 0.002% SDS, indicating that the presence of SDS at these concentrations completely suppressed the EP-QuIC reaction.

The nature of the recPrP substrate plays a crucial role in the RT-QuIC reaction, as different recPrP fragments exhibit varying seeding efficiencies when tested with the same CSF samples [33]. In our analysis, human recPrPs (recPrP 23–231 and recPrP 90–231) outperformed hamster recPrP 90–231 in terms of reaction efficiency (Supplementary Fig. 1C). In addition, hamster recPrP supported strong amplification at brain homogenate concentrations of 10^−6^ to 10^−9^ g; however, its reactivity declined sharply at lower concentrations. In contrast, both human recPrPs consistently produced robust ThT fluorescence signals at concentrations of 10^−11^ g, indicating substantially higher sensitivity of EP-QuIC for detecting minimal prion seeding activity. These findings suggested that the most effective EP-QuIC conditions included using human recPrP 23–231 or 90–231, incubation at 42°C, and exclusion of SDS from the reaction mix.

#### EP-QuIC enables the detection of PrP^Sc^ in CSF from sCJD patients

Using the optimized EP-QuIC protocol with human recPrP 23–231 as the substrate, we analyzed CSF samples from P3–P12 (Supplementary Table 1). All samples from definitively diagnosed cases tested positive within 6 h, with some showing clearly detectable signals as early as 4 h. In contrast, all non-prion disease controls remained negative, including N1–N3 (patients with epilepsy or Alzheimer’s disease) and N4–N10 (patients with corticobasal degeneration, Pick’s disease, amyotrophic lateral sclerosis, dementia with Lewy bodies, or frontotemporal lobar degeneration) (Fig. 3 and Supplementary Fig. 2). These findings indicated that EP-QuIC enables rapid and highly sensitive detection of PrP^Sc^ in CSF, demonstrating its potential as a practical diagnostic tool for prion diseases.

**Figure 3.**
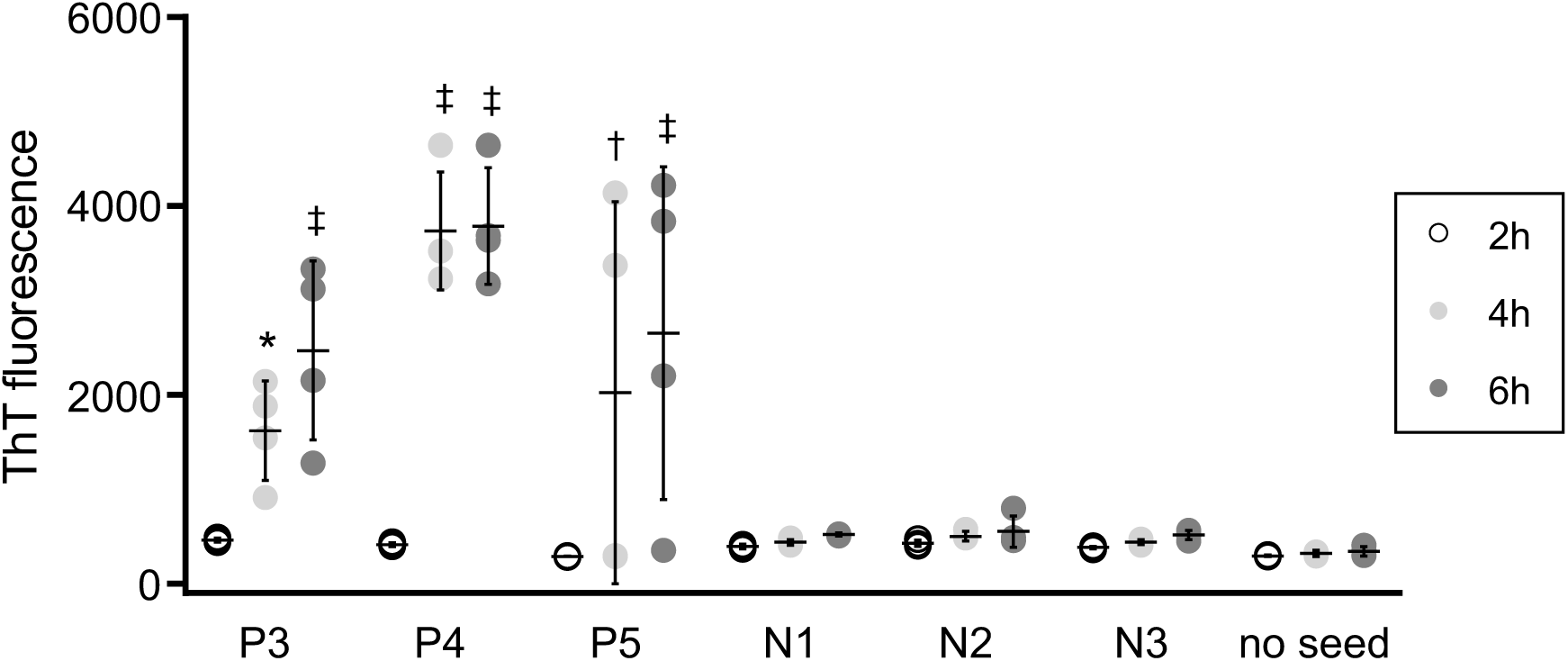
Application of EP-QuIC to cerebrospinal fluid (CSF) from patients with sCJD. CSF samples from patients with sCJD (P3–P5, n = 4 each) and non-prion disease controls (N1–N3, n = 4 each) were analyzed using EP-QuIC under optimized conditions, including the use of human recPrP 23–231 (batch #1), incubation at 42°C, and exclusion of SDS. ThT fluorescence was measured at 2, 4, and 6 h. All sCJD samples yielded positive ThT fluorescence signals within 6 h, whereas all control samples and “no-seed” reactions remained negative. \**P* < 0.05, †*p* < 0.01, ‡*p* < 0.001 *vs.* the “no-seed” controls at the corresponding time points.

## Discussion

In this study, we applied the EP-QuIC method using the Eppendorf Thermomixer^®^ C with minor modifications to assay conditions and evaluated its performance and compared it with that of conventional RT-QuIC. The EP-QuIC reactions were performed at 42°C, and the assay enabled rapid detection of prion seeding activity in the brain tissues from P1 and P2, with positive signals observed within 6 h—and, in some cases, as early as 2 h (Fig. 1). Notably, no increase in fluorescence was observed for “no-seed” controls, indicating the absence of spontaneous fibril formation under these conditions. EP-QuIC assay exhibited high sensitivity, with SD_50_ values from brain tissues comparable to those obtained from RT-QuIC performed over 24 h (Table 1). This finding indicated that EP-QuIC achieves sensitivity equivalent to that of RT-QuIC in a significantly shorter timeframe and without requiring high-performance plate readers with programmable shaking and temperature control.

Several studies have attempted minor optimizations, including changes in buffer composition, SDS concentration, and shaking speed; however, fundamental issues—such as extended assay duration, mechanical instability, and recPrP batch variability—remain unresolved. Notably, spontaneous fibril formation and poor reproducibility caused by batch-to-batch variability are persistent challenges that complicate the clinical application of RT-QuIC [26–28]. The modified EP-QuIC protocol developed in this study also exhibited improved reproducibility across different recPrP batches (Fig. 2). With RT-QuIC, certain human recPrP batches (#2 and #3) induced spontaneous ThT signal increases in unseeded reactions, impairing assay reliability. In contrast, EP-QuIC reactions using the same batches exhibited no detectable signal in unseeded controls. Furthermore, EP-QuIC reactions seeded with brain tissues from batch #2 displayed a clear signal onset at concentrations of 10^−7^–10^−9^ g within 6 h, whereas those from batch #3 yielded detectable signals at concentrations of 10^−7^–10^−8^ g. These results suggested that EP-QuIC is more tolerant to substrate variability and provides more consistent results. We also evaluated the influence of SDS, a known modulator of RT-QuIC sensitivity, on EP-QuIC. Although 0.001%–0.002% SDS has been reported to enhance reactivity in RT-QuIC [29,32], EP-QuIC reactions were completely suppressed at both SDS concentrations (Supplementary Fig. 1). This inhibition might stem from SDS-induced foaming under vigorous shaking conditions. Similarly, although incubation at 55°C allowed rapid signal detection within 2 h, spontaneous aggregation became prominent by 6 h, confounding interpretation. These findings underscored the use of 42°C temperature and omission of SDS as optimal conditions for EP-QuIC using human recPrP 23–231, with endpoint measurement at 4–6 h. Under these optimized conditions, EP-QuIC successfully distinguished CSF samples from patients with CJDs (P3–P12) and ten non-prion controls (N1–N10), achieving 100% sensitivity and specificity within 6 h (Fig. 3 and Supplementary Fig. 2). This finding demonstrated the potential of EP-QuIC as a rapid and reliable diagnostic tool for prion diseases. Furthermore, we retrospectively analyzed brain tissues from two previously reported sCJD cases using the EP-QuIC protocol. Although the sample size was limited, our findings supported the assay’s applicability across specimen types. Further statistical validation using larger cohorts will be necessary to establish robust cutoff thresholds for routine clinical use.

A fundamental distinction between EP-QuIC and RT-QuIC lies in their methods of fluorescence data acquisition. RT-QuIC captures fluorescence signals in real time, allowing kinetic analysis that helps differentiate early true-positive signals from late-arising false positives. This technique typically requires continuous use of fluorescence plate readers such as the Infinite® 200 PRO or FLUOstar Omega®, which operate under prolonged high-speed orbital shaking (432–700 rpm) with alternating agitation and rest cycles at controlled temperatures. However, this setup ties up a single fluorescence plate reader for the entire duration of the assay and subjects the instrument to significant mechanical stress, often resulting in equipment wear, higher maintenance demands, and increased operational costs. In contrast, EP-QuIC capitalizes on the mechanical durability and thermal precision of thermomixers, which provide consistent high-speed shaking and precise temperature control. These instruments are mechanically robust, user-friendly, and cost-effective, without the need for integrated optics or proprietary software. This feature enables parallel processing of multiple plates. Fluorescence measurements are performed only once at the endpoint using a standard plate reader, significantly increasing throughput and reducing instrumentation costs. The primary limitation of EP-QuIC is its inability to monitor PrP^Sc^ formation in real time. Instead, it relies on endpoint fluorescence detection, which—when properly validated—offers sufficient resolution to distinguish positive from negative samples under optimized assay conditions. This characteristic enables more flexible, scalable, and cost-efficient workflows, especially in environments where real-time kinetic tracking is not required. Although the mechanisms underlying the enhanced speed and sensitivity of EP-QuIC remain unclear, they present a promising direction for future research.

In conclusion, EP-QuIC represents a fast, reliable, and user-friendly amplification method that overcomes key limitations of RT-QuIC, including its reliance on specialized equipment and susceptibility to recPrP batch variability. With future enhancements—such as the use of PrP fibril-specific antibodies or the adoption of colorimetric detection—and potential integration with wire-QuIC for assessing surface decontamination, EP-QuIC can exhibit great potential to become a versatile platform. Beyond prion disease diagnostics, it might also be adapted for broader applications in other protein misfolding disorders, such as α-synucleinopathies.

## Data Availability

All data generated or analyzed during this study are included in the manuscript and its supplementary information files.

## Acknowledgments

We thank Kazunori Sano, Yuzuru Taguchi, Kaori Ono-Ubagai, and Hiroya Tange from Nagasaki University for helpful discussions and critical assessment of the manuscript, and Atsuko Matsuo, Hanako Nakayama, Marie Yamaguchi, and Megumi Tanaka for technical assistance. Finally, we thank Enago (www.enago.jp) for editing this manuscript.

## Competing interests

The authors declare no competing interests.

**Supplementary** Figure 1**. Effect of temperature, sodium dodecyl sulfate SDS, and substrate on EP-QuIC efficiency.** A) Effect of temperature on EP-QuIC kinetics. Reactions were performed at 55°C using human recPrP 23–231 (batches #1, #2, and #3) and brain homogenates from a patient with sCJD (P1). ThT fluorescence was measured at 2, 4, and 6 h. \**P* < 0.05, †*p* < 0.01, ‡*p* < 0.001 *vs.* the “no-seed” controls at the corresponding time point. B) Effect of SDS on EP-QuIC. Reactions were conducted at 42°C with 0%, 0.001%, or 0.002% SDS using human recPrP 23–231 (batch #1) and brain homogenates from a patient with sCJD (P1). ThT fluorescence was measured at 2 and 4 h. †*P* < 0.01, ‡*p* < 0.001 *vs.* the “no-seed controls” at the corresponding time points. C) Comparison of different recPrP substrates. Human recPrP 23–231 (batch #1), human recPrP 90–231, and hamster recPrP 90–231 were evaluated at 42°C with brain homogenates from a patient with sCJD (P1). ThT fluorescence was measured at 2, 4, and 6 h. †*P* < 0.01, ‡*p* < 0.001 *vs.* the “no-seed controls” at the corresponding time points.

**Supplementary** Figure 2**. Application of EP-QuIC to CSF from non-prion disease controls.** CSF samples from non-prion disease controls (N4–N10, n = 4 each), from the patients with CJDs (P6–P12, n = 4 each), brain homogenates from a patient with sCJD (P1) at 10^−7^ g, and artificial cerebrospinal fluid (A-CSF) were analyzed using EP-QuIC under optimized conditions, including the use of human recPrP 23–231 (batch #1), incubation at 42°C, and exclusion of SDS. ThT fluorescence was measured at 2, 4, 6, and 8 h. The homogenates yielded positive ThT fluorescence signals, whereas all control samples, A-CSF, and “no-seed” reactions remained negative. \**P* < 0.05, †*P* < 0.01, ‡*p* < 0.001 *vs.* the “no-seed” controls at the corresponding time points.

