## Supplementary figures and images for "Optimization of a thermal shaker-aided quaking-induced conversion platform for the rapid detection of prion seeding activity in human samples"

### Supplemetary Figure 1

#### Supplementary Fig.1

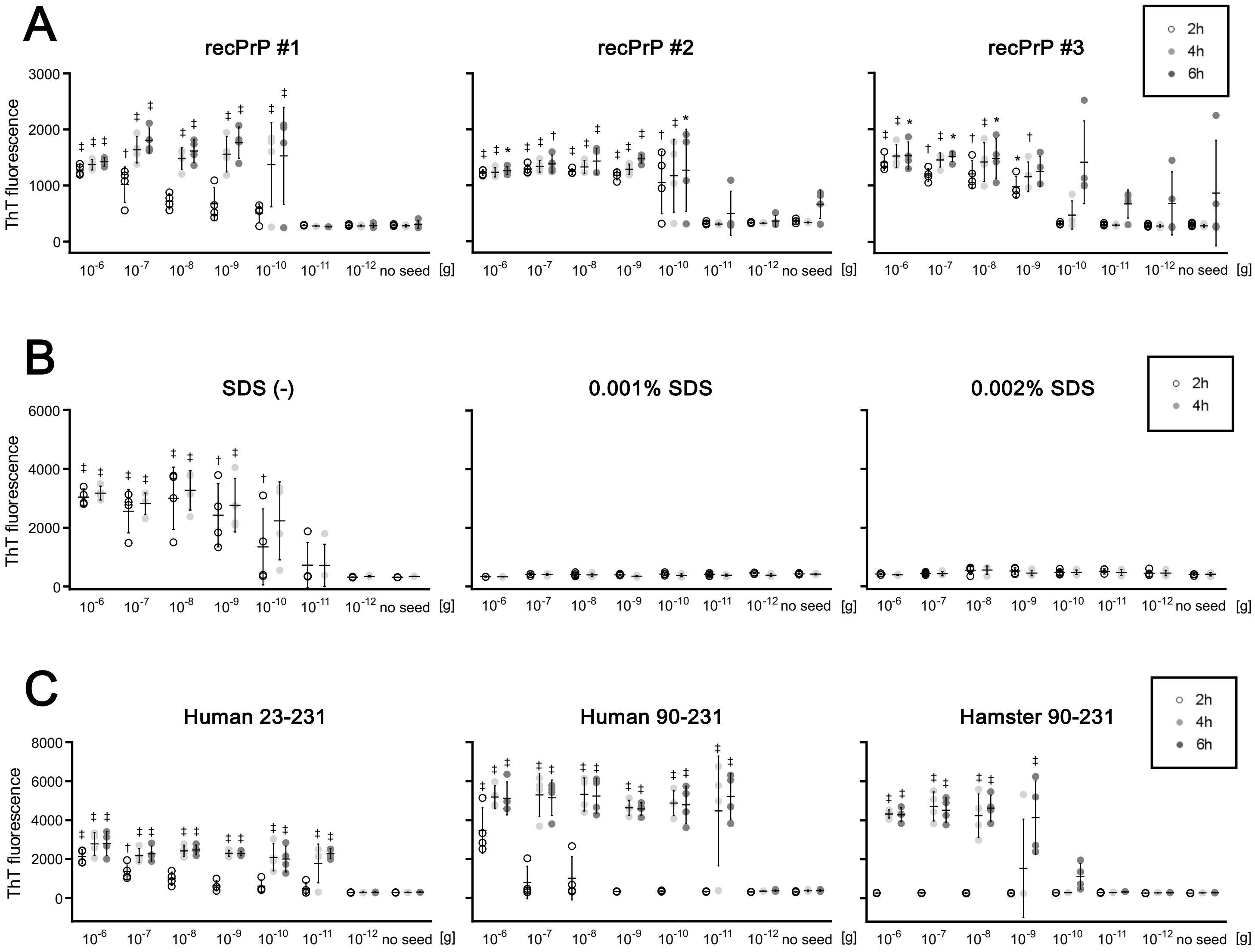

### Supplemetary Figure 2

#### Supplementary Fig.2

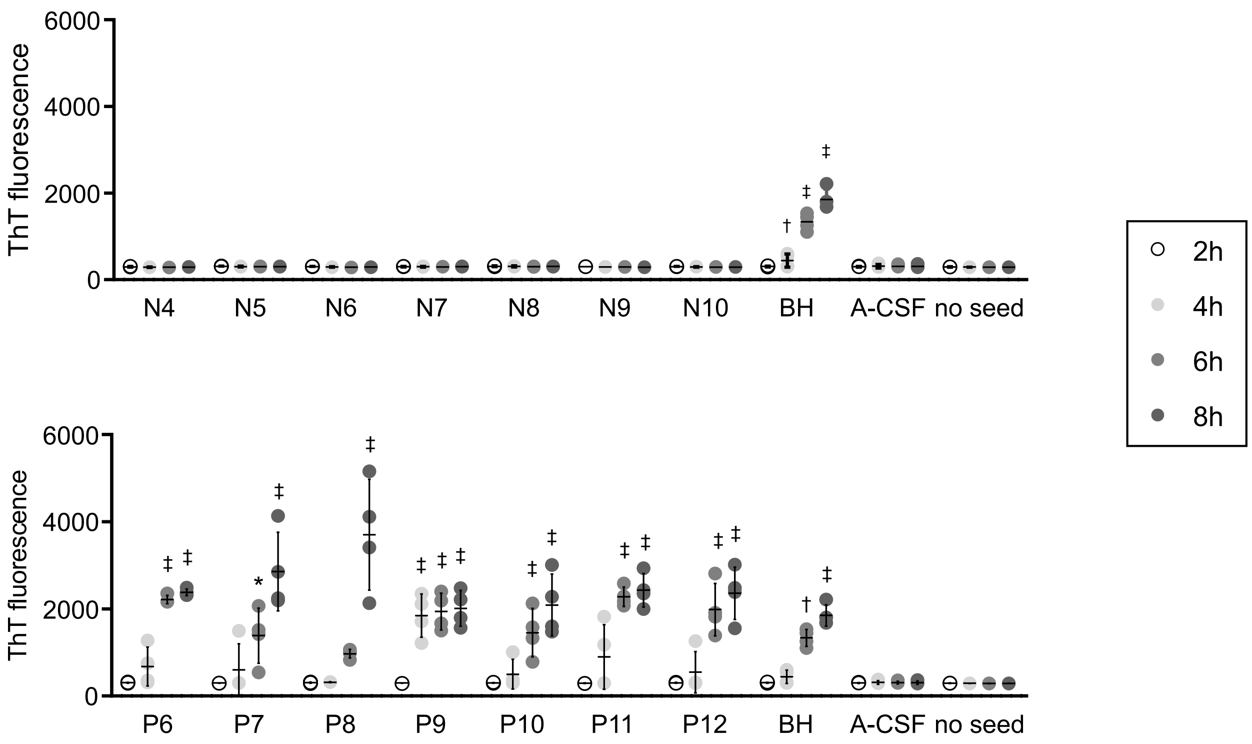
