## Supplementary material for "Optimization of a thermal shaker-aided quaking-induced conversion platform for the rapid detection of prion seeding activity in human samples": Supplemetary Table 1

**Supplementary Table 1.** Demographic and clinical characteristics of sCJD patients (P3–P12) and non-prion disease controls (N1–N10) whose CSF samples were analyzed in this study.

| **ID** | **Sex** | **Age range** | **Definitive diagnosis** |
|---|---|---|---|
| P3 | Female | 81–85 | sCJD |
| P4 | Male | 71–75 | sCJD |
| P5 | Female | 66–70 | sCJD |
| P6 | Female | 66–70 | gCJD (E200K) |
| P7 | Female | 71–75 | sCJD |
| P8 | Female | 71–75 | sCJD |
| P9 | Female | 81–85 | sCJD |
| P10 | Female | 71–75 | sCJD |
| P11 | Male | 81–85 | sCJD |
| P12 | Female | 76–80 | sCJD |
| N1 | Female | 76–80 | Epilepsy |
| N2 | Male | 81–85 | Epilepsy |
| N3 | Female | 56–60 | Alzheimer’s disease |
| N4 | Female | 66–70 | Corticobasal degeneration |
| N5 | Female | 71–75 | Corticobasal degeneration |
| N6 | Female | 86–90 | Pick's disease |
| N7 | Female | 66–70 | Amyotrophic lateral sclerosis |
| N8 | Male | 81–85 | Dementia with Lewy bodies |
| N9 | Female | 86–90 | Dementia with Lewy bodies |
| N10 | Male | 76–80 | Frontotemporal lobar degeneration with TDP-43 inclusions |
